# Metabolomic Changes in Idiopathic and *GBA1* Parkinson’s Disease

**DOI:** 10.1101/2025.09.08.25335343

**Authors:** Caitlin Walton-Doyle, Elisa Menozzi, Anthony H.V. Schapira, Perdita Barran

## Abstract

**Background:** Variants in the β-glucocerebrosidase (*GBA1*) gene are the commonest genetic risk factor for Parkinson Disease (PD). Here, we use mass spectrometry based metabolomics to analyse serum and sebum samples from 50 genotyped participants and find differences in lipid and sugar regulation, oxidative stress and the production of amino acids and neurotransmitters which distinguish *GBA1*-PD from iPD. A subset of highly ranked features also correlate to *GBA1* variant severity.

**Methods:** Randomised and blinded serum samples from *GBA1*-PD and iPD participants (*n*=50) were analysed by Liquid Chromatography – Mass Spectrometry (LC-MS) and Gas Chromatography – Mass Spectrometry (GC-MS), and sebum samples by headspace GC-MS. After deconvolution and alignment of data, three models were created: *GBA1*-PD *vs*. iPD, severity of *GBA1* variant, and drug naïve *vs.* medicated.

**Findings:** Differences in metabolomic signatures were seen between *GBA1*-PD and iPD in sebum and serum with good specificity and sensitivity. Significant pathways in serum included sphingolipid metabolism, amino sugar metabolism and amino acid pathways, whereas significant features between groups in sebum are hypothesised to be lipid degradation products. Several highly ranked features displayed regulation with variant severity. When separating participants according to medication status, we see separation by supervised analysis by all analytical techniques.

**Interpretation:** Significant pathways and metabolites between *GBA1*-PD and iPD display alterations in sphingolipids. Other features and pathways that may be related include amino sugar metabolism and lipid breakdown products in sebum. Together, significant features indicate possible changes in mechanisms of oxidative stress and neurotransmitter precursors.

**Funding:** AHVS thanks the Kattan Trust and Cure Parkinson’s for their generous support of this research. PEB thanks the Michael J Fox Foundation (grant ref:12921) BBSRC (grant Ref BB/L015048/1) and Parkinson’s UK (grant ref: K-1504) for funding our work.

**Research In context:** *Evidence before this study:* Variants in the *GBA1* gene are a genetic risk factor for PD, associated with earlier onset and faster progression. *GBA1* encodes GCase, and variants can lead to accumulation of glucosylceramide and other substrates. Prior metabolomics studies have found alterations in amino acids (Greuel) and lipids (Guedes) and have noted the complexity of relating glycosphingolipid levels to mutation (den Heijer). Only a handful of studies have investigated variation with mutation severity, and those that exist focus on the presence of proteins such as alpha synuclein or GCase. Further, non-invasively sampled biofluids such as sebum have not been investigated for metabolomic signatures and remain underexplored as sources of biomarkers.

*Added Value of this study:* This study integrates serum metabolomics with headspace profiling of sebum. Findings reveal differences in lipid and sugar regulation, amino acid pathways and oxidative stress mechanisms. Both serum and sebum reveal differential profiles of *GBA1*-PD and iPD, as well as containing significant features that display regulation with gene severity.

*Implications of all the available evidence:* The work demonstrates that *GBA1* variants and their severity alter the metabolomic profiles of PD, which can be readout from both serum and sebum. Quantifying the metabolic signatures of differentially dysregulated pathways could be used to target therapeutics for genetically defined PD subtypes and improve understanding of the impacts of gene mutations on disease progression. Graphical Abstract

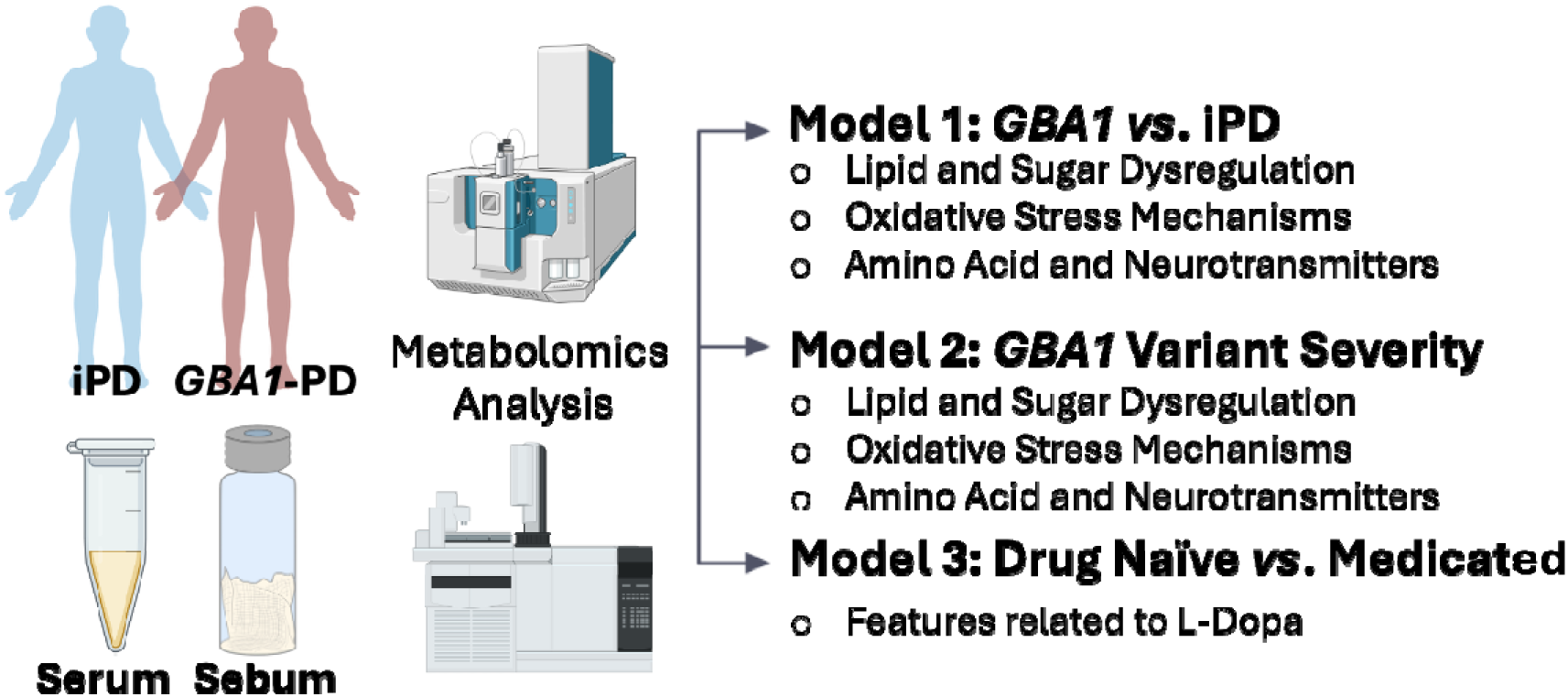

## Introduction

Variants in the β-glucocerebrosidase (*GBA1*) gene occur in up to 15% of Parkinson’s Disease (PD) patients.^1^ The *GBA1* gene encodes the lysosomal enzyme glucocerebrosidase (GCase) which cleaves the sphingolipid glucosylceramide (GlcCer) into glucose and ceramide.^2^ Bi-allelic pathogenic variants in *GBA1* cause Gaucher disease (GD), characterised by intralysosomal accumulation of GlcCer and its deacylated form glucosylsphingosine (GlcSph) in cells, particularly macrophages. This results in clinical manifestations such as anaemia, thrombocytopenia, hepatosplenomegaly and bone involvement. Depending on the absence or presence of neurological manifestations in GD patients, variants in *GBA1* are classified as mild or severe, the latter being associated with a much more aggressive form of GD.^2^ Within PD, the classification of *GBA1* variants into mild and severe based on GD severity is maintained, while variants that do not cause GD but have been found to be more prevalent in PD than controls are called risk variants.^2^ Clinically, some genotype-phenotype correlations have been reported, with severe variant carriers presenting the more rapidly progressing phenotype,^3^ but unequivocal markers of disease penetrance, severity and progression are lacking. Several mechanisms have been implicated in *GBA1*-PD pathogenesis, including alterations in the autophagy system, endoplasmic reticulum (ER) stress, mitochondria dysfunction and dysregulation in the lipid membrane composition.^2^ Understanding these processes and translating them into biomarkers which are clinically relevant and minimally invasive for the patients, remain a challenge.

Serum metabolites have displayed alterations in PD, including perturbations in amino acid metabolism and oxidative stress markers.^4^ Sebum, the oily secretion from sebaceous glands, has shown metabolomic differences between PD patients and controls, including lipids and products of lipid degradation as well as differential profiles for prodromal symptoms.^5–9^ This discrimination has shown the great potential of sebum as a non-invasive biofluid, with possibility of at-home sampling.^10^ Analysis has previously been implemented to compare idiopathic PD patients (i.e., without known genetic predisposition, iPD) and patients with genetic predisposition (e.g., carriers of *GBA1* variants, *GBA1*-PD). For instance, plasma metabolomics has revealed increased glutamine, glycine and other amino acids in *GBA1*-PD against iPD, and lipidomic analysis has found increased ceramide and decreased phosphatidylethanolamine in *GBA1*-PD.^11,12^ Investigation into metabolomic features as a function of severity of *GBA1* variant has not been previously undertaken.

We investigated metabolomic differences in idiopathic PD (*n*=25), with confirmed wild-type *GBA1* status and PD participants with heterozygous variants in *GBA1* (*GBA1*-PD, n=25), representative of severe, mild and risk variant types. We analysed serum by Gas Chromatography – Mass Spectrometry and Liquid Chromatography – Mass Spectrometry as well as the volatile molecules of sebum by Headspace GC-MS. Both biofluids showed discrimination between the two phenotypes by supervised, multivariate analysis, with serum identifying many pathways as significant including sphingolipid metabolism, amino sugar metabolism, and multiple amino acid pathways, and significant features in sebum being annotated as lipid degradation products, replicating our previous findings.^6,7^ We subsequently investigated the altered metabolomes with severity of *GBA1* variants and probed features highly ranked in classification of *GBA1*-PD against iPD which displayed regulation over severity. Putative identifications for features regulated with severity were assigned as fatty acyls, sphingolipids, glycerophospholipids, lactate metabolites and dopamine pathway metabolites. Finally, we investigated differences between participants medicated for PD against those who were not (drug naïve) regardless of their genetic status, and identified significant features between these groups, several of which were directly related to dopamine.

## Methods

Participants were recruited via the RAPSODI GD and PD Frontline study, from University College London (UCL).^13^ This study was approved by the local Ethics Committees (London – Queen Square REC: 15/LO/1155). All participants signed informed consent upon enrolment. The analysis of the *GBA1* gene was performed using saliva samples.^14^

The work was split into three models, to examine differences between iPD and *GBA1*-PD, to examine alterations as a function of *GBA1* variant severity, and finally to probe changes upon administration of PD medication. The samples included in each model are listed below in Table 1 and demographics are listed in Supplementary Information, Table S 1. Materials, methods and data analysis are described in sections SI 1 and SI 2.

**Table 1:** Samples included in each of the models. For serum, three analytical techniques were undertaken on 50 samples, and for sebum, headspace GC-MS was performed on 50 samples from the same participants with 2 failed injections.

| **Model** | **Participants Serum** | **Participants Sebum** |
| --- | --- | --- |
| *GBA1-PD vs. iPD* | 25 vs. 25 | 24 vs. 24 |
| *GBA1 Severity* | 12 risk vs. 6 mild vs. 7 severe | 12 risk vs. 6 mild vs. 6 severe |
| *Medication* | 35 vs. 15 | 33 vs. 15 |
| *Analysis Type* | LC-MS Positive ionisation mode,<br>LC-MS Negative ionisation mode,<br>Liquid injection GC-MS | Headspace GC-MS |

## Results

### Model 1: *GBA1*-PD *vs*. iPD

Serum features ranked highly in the separation of *GBA1*-PD and iPD by LC-MS and GC-MS were inputted to ROC multivariate analysis, displayed in igure 1A. The resultant ROC displayed good specificity and sensitivity (AUC=0.932 with 100 features). By both polarities in LC-MS the two groups separated completely by supervised scores plots (Figure S 1 A and B). Pathway analysis was performed through Metaboanalyst ^15^ using Mummichog, ranked by t score and annotated by the homo sapiens KEGG library. Pathways found to be significantly different between the two groups are listed below in Table 2.

**Table 2:**
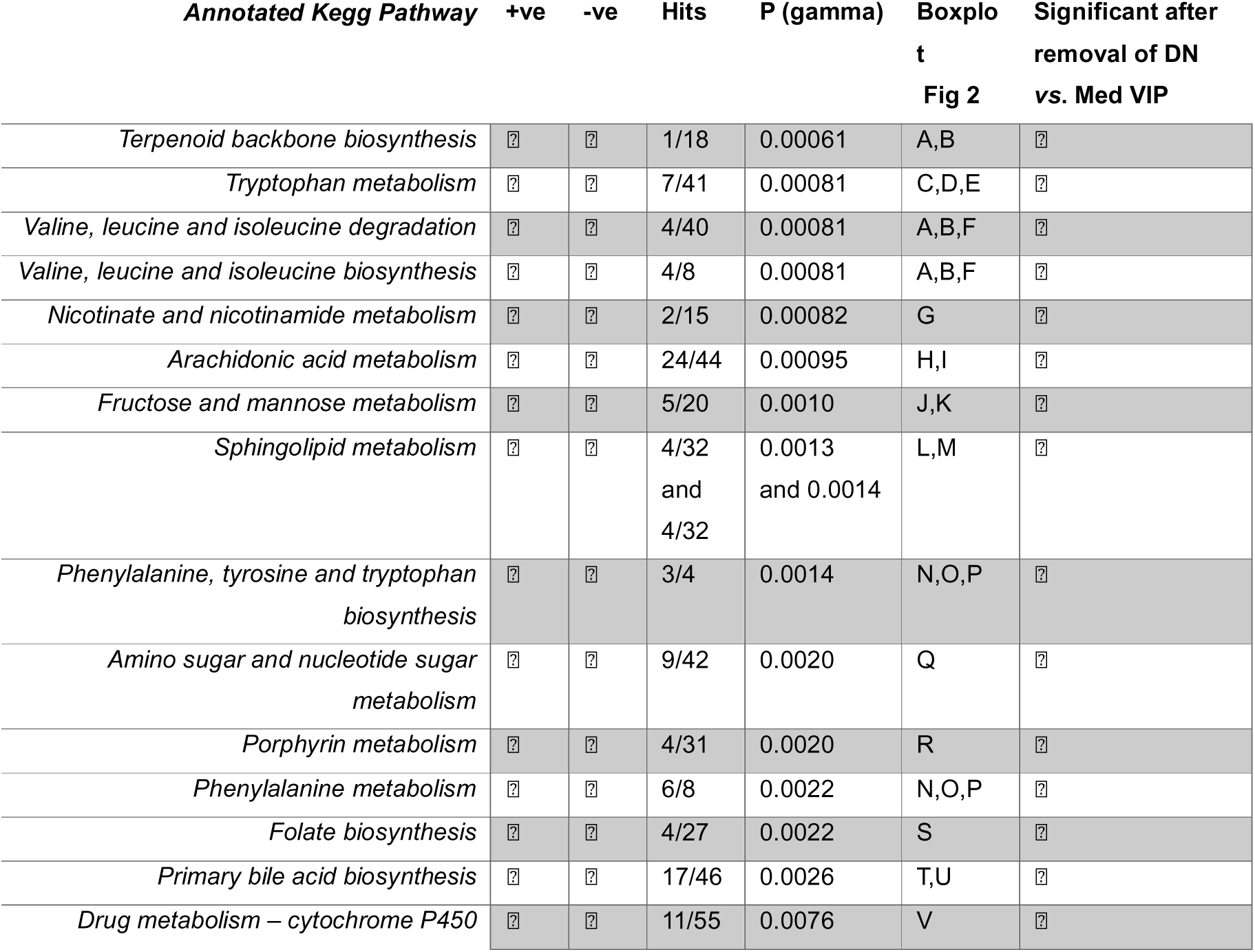
Mummichog pathway analysis of samples analysed by LC-MS in positive and negative ionisation comparing phenotypes GBA1-PD and iPD. Fifteen pathways were identified as significant, with one seen in both polarities. The hits here are features assigned to metabolic networks. To ensure the features were not a result of medication administered, the highly ranked features from a drug naïve vs. medicated model were removed, and most remained significant.

| **Annotated Kegg Pathway** | **+ve** | **-ve** | **Hits** | **P (gamma)** | **Boxplot**<br>**Fig 2** | **Significant after**<br>**removal of DN**<br>**vs. Med VIP** |
| --- | --- | --- | --- | --- | --- | --- |
| *Terpenoid backbone biosynthesis* | ? | ? | 1/18 | 0.00061 | A,B | ? |
| *Tryptophan metabolism* | ? | ? | 7/41 | 0.00081 | C,D,E | ? |
| *Valine, leucine and isoleucine degradation* | ? | ? | 4/40 | 0.00081 | A,B,F | ? |
| *Valine, leucine and isoleucine biosynthesis* | ? | ? | 4/8 | 0.00081 | A,B,F | ? |
| *Nicotinate and nicotinamide metabolism* | ? | ? | 2/15 | 0.00082 | G | ? |
| *Arachidonic acid metabolism* | ? | ? | 24/44 | 0.00095 | H,I | ? |
| *Fructose and mannose metabolism* | ? | ? | 5/20 | 0.0010 | J,K | ? |
| *Sphingolipid metabolism* | ? | ? | 4/32<br>and<br>4/32 | 0.0013<br>and 0.0014 | L,M | ? |
| *Phenylalanine, tyrosine and tryptophan*<br>*biosynthesis* | ? | ? | 3/4 | 0.0014 | N,O,P | ? |
| *Amino sugar and nucleotide sugar*<br>*metabolism* | ? | ? | 9/42 | 0.0020 | Q | ? |
| *Porphyrin metabolism* | ? | ? | 4/31 | 0.0020 | R | ? |
| *Phenylalanine metabolism* | ? | ? | 6/8 | 0.0022 | N,O,P | ? |
| *Folate biosynthesis* | ? | ? | 4/27 | 0.0022 | S | ? |
| *Primary bile acid biosynthesis* | ? | ? | 17/46 | 0.0026 | T,U | ? |
| *Drug metabolism – cytochrome P450* | ? | ? | 11/55 | 0.0076 | V | ? |

The features contributing to the pathways were compared between the two phenotypes. Some pathways were annotated with multiple features differing in regulation, likely due to upregulation and accumulation of a feature in one part of the network repressing another part. Box and whisker plots of the features of the named pathways are displayed below in Figure 2A-V and a full list of pathways and regulation is listed in Supplementary Information Table S2

The GC-MS serum displayed discrimination but not complete separation between the two groups by PLS-DA scores plot ((Figure S1 C). Of the 365 robust features, 51 had a VIP score > 1 in PC1. Pathway analysis was undertaken using the identifications of these features, with only the tyrosine metabolism pathway determined as significant (*p* = 0.0097 Figure S2). Two features contributed to the pathway and were annotated as dopamine and 3,4-dihydroxy-L-Phenylalanine, more commonly known as l-DOPA shown in Figure 2W and X. Dopamine was upregulated in the *GBA1*-PD cohort whereas l-DOPA displayed higher intensity in the iPD cohort. Investigation was done to ensure significant pathways were not due to medication (as more participants in iPD were drug naïve compared to *GBA1*-PD). Pearson’s correlation was undertaken for all features against medication (SI 3) and no features contributing to the pathways were found significant (correlation ≥0.8 or ≤-0.8 and significance <0.05). Features that had a VIP score > 1 in the drug naïve *vs*. medicated model (described below) were then removed from the matrix and pathway analysis was recalculated. Pathways not found significant after removal of these features and thus may be related to medication, are labelled in Table 2. By GC-MS the tyrosine pathway was no longer found as significant, and for this reason, it can be inferred the metabolites could be related to medication.

The sebum samples *GBA1*-PD (*n* = 24) and iPD (*n* = 24) show discrimination by supervised scores plot but the groups could not be separated fully (Figure S1 D), similar to the GC-MS serum analysis. Of the 535 features, 118 had a VIP score > 1. The VIP features were used as the input of a multivariate ROC curve (Figure 1B) displaying good sensitivity and specificity (AUC =0.832 with 100 features). Only thirteen VIP features could be assigned putative identifications: five alkanes, seven fatty acid methyl esters (FAMEs) and one fatty acid (FA) derivative, the box plots are displayed in Figure 3. We have previously evidenced these are likely products of degradation of larger molecules.^6^

**Figure 1:**
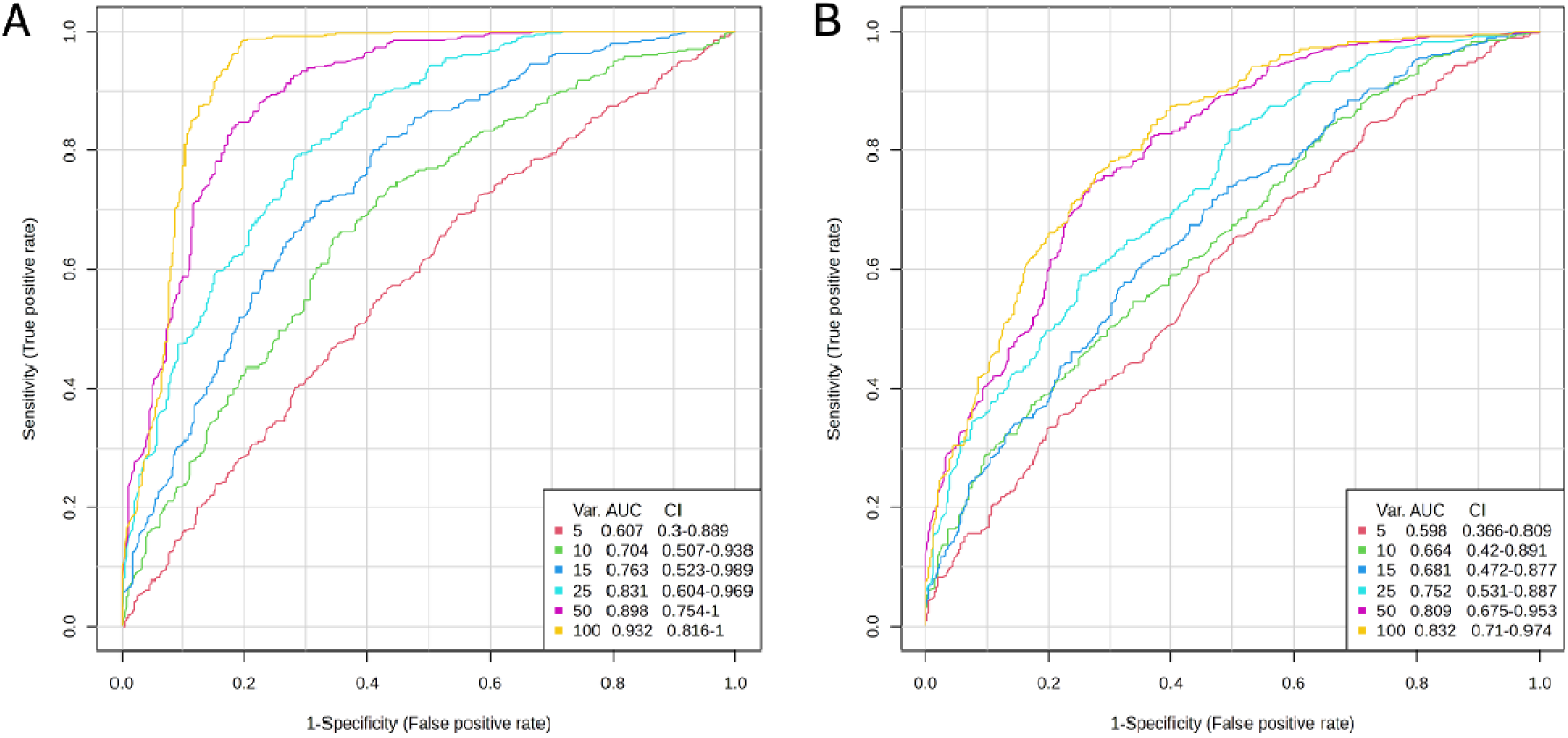
Multivariate ROC curves of GBA1-PD vs. iPD, A) highly ranked VIP features from the three serum analyses and B) highly ranked VIP features from the Headspace sebum analysis. ROC curves were generated with Monte-Carlo Cross Validations (MCCV) using 2/3 samples to train and 1/3 to validate. Linear SVM was the classification and ranking method.

**Figure 2:**
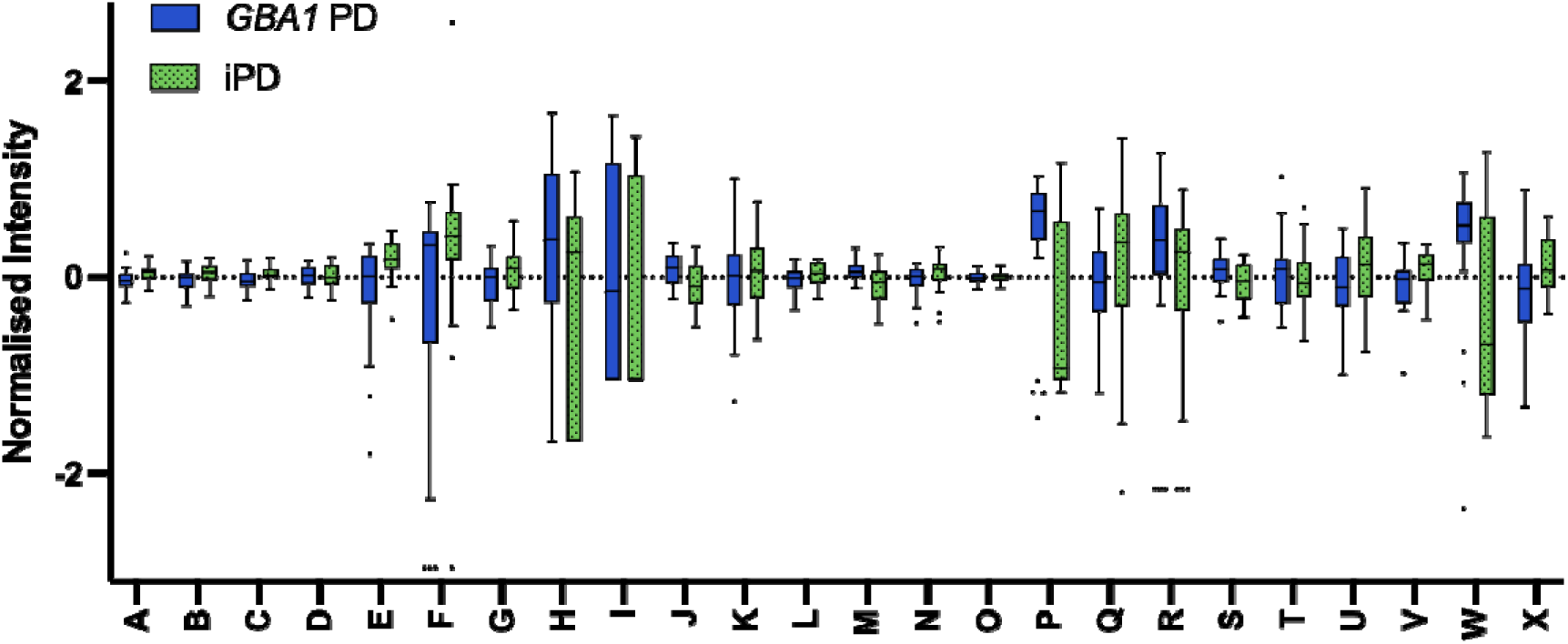
Box and whisker plots of features of serum contributing to pathways with significance between the GBA1-PD (blue) and idiopathic PD (green) phenotypes. The pathways are listed in Table 2, with the addition of feature W and X (found significant by GC-MS) as dopamine and 3,4-Dihydroxy-L-phenylalanine respectively.

**Figure 3:**
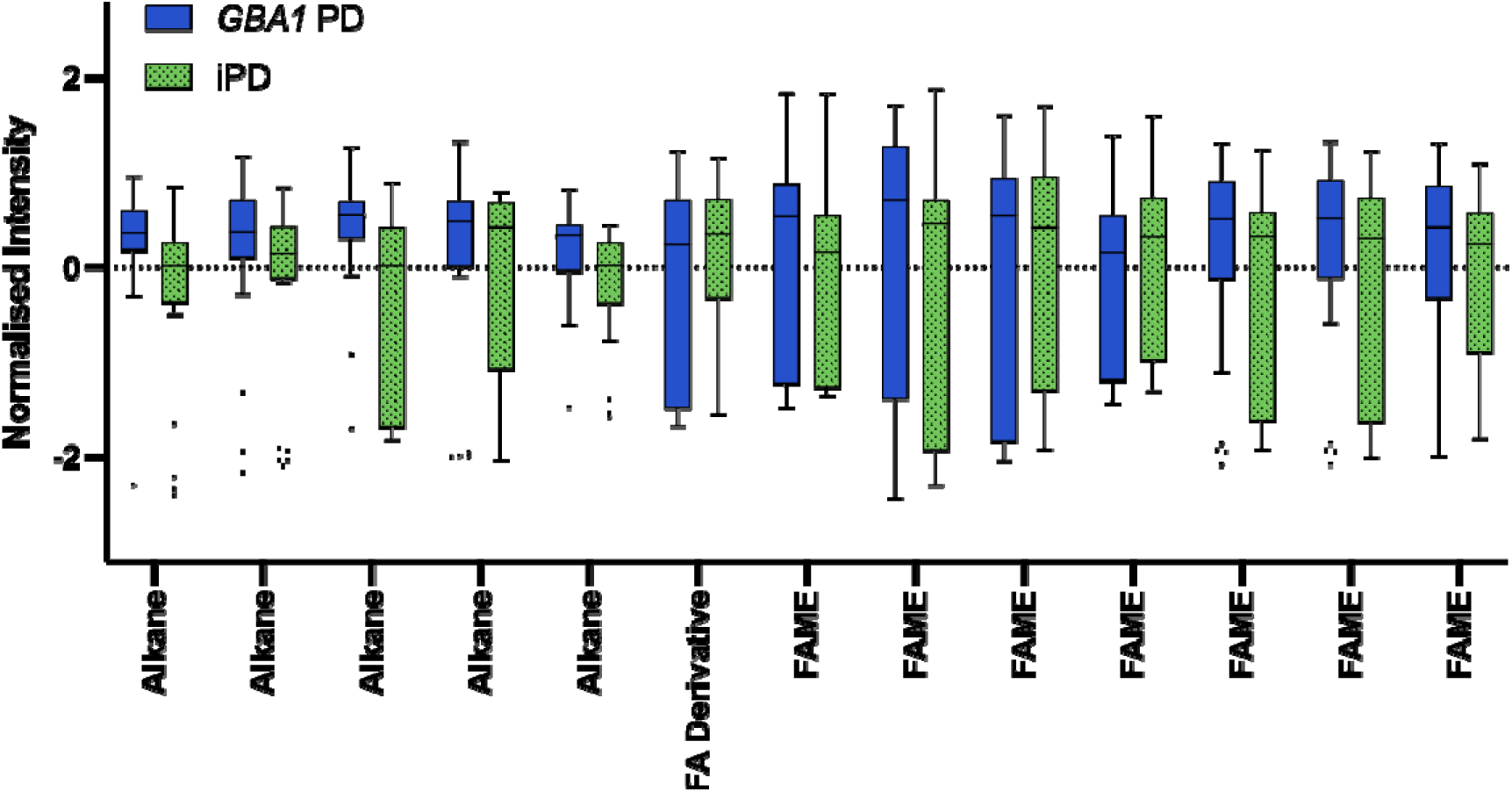
Box and whisker plots of sebum features that had VIP score >1 in the PLS-DA of GBA1-PD (blue) vs. iPD (green) and could be assigned putative identifications. A fatty acid derivative and a FAME were upregulated in iPD whereas five alkanes and six FAMEs were upregulated in GBA1-PD.

### *GBA1* variant severity

The participants with *GBA1* variants were separated according to variant severity,^16^ namely: risk (*n* = 12), mild (*n* = 6) and severe (*n* = 7). This was first modelled using PLS-DA where scores plots showed separation with small amounts of overlap, but, likely due to the smaller numbers of samples the models could not be validated. Following this, the features that were highly ranked from the *GBA1*-PD *vs*. iPD model were investigated as a function of severity. For both ionisation modes of LC-MS, the top 50 VIP features were taken, for GC-MS, all VIP > 1 were used (51 for serum and 118 for sebum).

The LC-MS analysis revealed several features differentially regulated across severity with the intensity of the mild variant between that of risk and severe (16 in positive ionisation and 13 with negative ionisation). Of these features, 18 were assigned putative identifications, displayed in Figure 4.

**Figure 4:**
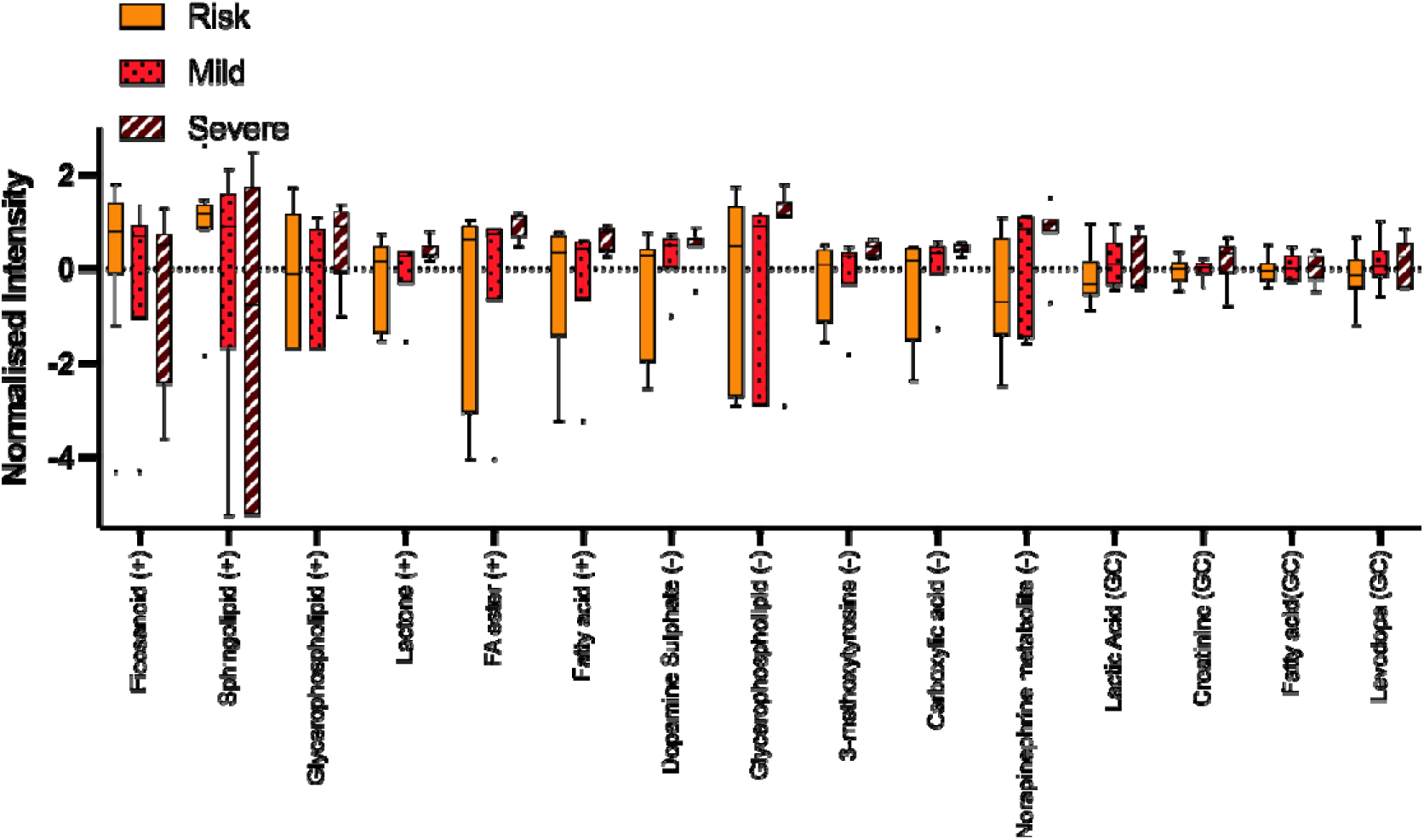
Box plots of features showing regulation with GBA severity, namely risk (orange), mild (red) and severe (maroon). In LC-MS positive ionisation (+) features show different regulation whereas by negative ionisation (-) and GC-MS, the features all decrease with severity.

Features identified as endogenous and of relevance include, an eicosanoid, a lactone, a FA, a FA ester, a sphingolipid, two glycerophospholipids, a tyrosine derivative, a carboxylic acid derivative, a metabolite of norepinephrine and a dopamine derivative in the form of dopamine sulphate. A dopamine precursor was also found to differentiate severity by the GC-MS serum analysis and showed a similar trend of increasing with severity.

Of the 51 features in the GC-MS serum analysis, 21 showed regulation, indicating these are possibly indicative of *GBA1* severity. Four could be assigned putative identifications of endogenous compounds as: lactic acid, creatinine, a FA and l-DOPA. Box and whisker plots are displayed in Figure 4. The pyruvate metabolism and glycolysis/ gluconeogenesis pathways were found significant, with lactic acid being the metabolite involved in both.

For the sebum analysis, of 118 features, 51 showed differential regulation over severity. Of these, four identifications were putatively assigned: an alkane (increasing with severity), two FAMEs (increasing with severity) and a fatty acid derivative (decreasing with severity).

### Dopaminergic drug use

The participants were separated into those administered medication for PD (medicated, *n*=35, 33 for sebum analysis) and those who had never been administered medication for PD (drug naïve *n*=15). PLS-DA scores plots were used to visualise differences between the classes and in all analyses, clustering was displayed (Figure S3).

Fold change (FC) and t tests were used to investigate significantly different features. Features with FDR corrected *p*-value < 0.05, and fold change threshold of 2 were considered significant. In LC-MS, positive ionisation gave eleven features and negative ionisation gave twelve, all upregulated in the medicated cohort.

Identifications of relevance were a lactone (+), a tyrosine derivative (both polarities), a fatty ester (+), a FA (+), a carboxylic acid derivative (-) and two dopamine derivatives (-). As tyrosine is a precursor to dopamine, it is hypothesised the tyrosine and dopamine derivatives are directly related to dopamine-based medication. The GC-MS analysis of serum gives just two metabolites as significant, both upregulated in the medicated cohort and one putatively assigned as dopamine. For sebum, though the groups separated by scores plot, no features were found to be significant. The features from all serum analyses are displayed in the volcano plot below (Figure 5).

**Figure 5:**
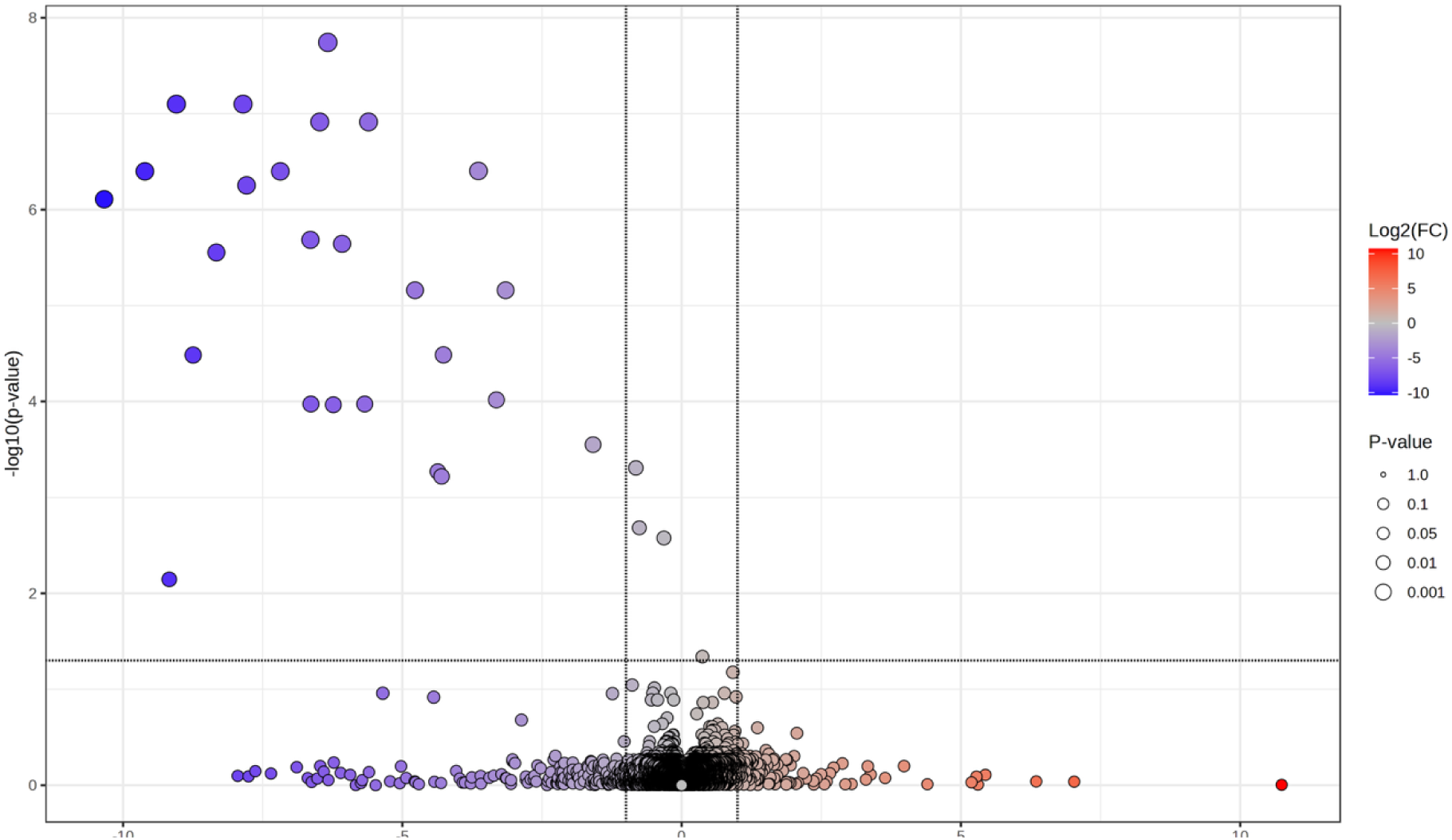
A volcano plot of the features in serum, showing significance between the drug naïve and medicated cohorts. Twenty-five features were found to have p-value <0.05 and fold change >2 by GC-MS and LC-MS, all of which were upregulated in the medicated cohort.

Pearson’s correlation was used to display relationships between medication and features (Section SI 3). Of the 25 features here, four showed correlation (correlation >0.8 and significance <0.05) with L-Dopa. The carboxylic acid derivative and a dopamine derivative were the only features of these four assigned an identification. The table is in Supplementary Information Table S4

Interestingly, in all serum analysis methods, there were features where five of the medicated individuals showed regulation away from the mean – in a similar range as the drug naïve individuals. On further investigation, these are the five medicated participants who are not taking levodopa (medications listed in Table S3). They are from both iPD and *GBA1*-PD groups, and are not the same gender, ethnicity or BMI category, thus it is reasonable to assume these features are directly correlated with levodopa. Example box and whisker plots to display this phenomenon are shown in Supplementary Information Figure S4.

## Discussion

Here, we have demonstrated metabolomic changes in serum and sebum between iPD and *GBA1*-PD, with mummichog analysis elucidating alterations in pathways between the groups. Highly ranked features were investigated as a function of severity of the gene variant, and a subset shows regulation with severity both in serum and sebum. Finally, comparing drug naïve against medicated participants displayed separation by scores plot, with some features showing correlation to administration of L-Dopa.

## Dysregulation of Lipids and Sugars

By LC-MS analysis of serum, a common pathway significantly altered in both polarities between *GBA1*-PD and iPD is sphingolipid metabolism, likely linked to GCase. A putative ID of sphingamine phosphate was upregulated in iPD whereas a feature identified as GluSph was upregulated in *GBA1*-PD, which has previously been displayed.^17^ In our work, we have previously reported an upregulation in sphingolipid metabolism between PD and control.^7^ As sphingolipids contain a fatty acid and hydrocarbon chain, the degradation of these larger lipid moieties may be detected as alkanes and fatty acids. It is plausible that such metabolites in the sebum headspace analysis are breakdown products related to lipid dysregulation, which we have previously demonstrated by headspace analysis of lipid standards.^6^ The presence of such from sebum analysis supports the general differential regulation found of several lipid pathways with LC-MS.

The amino and nucleotide sugar pathway dysregulation gives evidence for alterations in sugar metabolism. Glucose can be synthesised from nucleotide sugars, linking the pathway with GCase. The downregulation of the pathway in the *GBA1*-PD could suggest accumulation of amino sugars. Sugar structures have previously been associated with PD, where glycosaminoglycans are thought to be present in Lewy bodies and glycation of alpha synuclein associated with ease of oligomerisation.^18^ Previous lipidomics analysis of serum from PD patients classified as *GBA1* variant carriers *vs*. non *GBA1* carriers found decreased phosphatidylethanolamine (PE) among other lipid classes in the *GBA1* carrier group.^12^ A breakdown product of PE (glycerophosphoethanolamine) has been found in increased levels in putamen of *GBA1*-PD brains compared to controls.^19^ Here, when investigating with respect to variant severity, we complement these findings with identification of a glycerophospholipid increasing with severity of the variant, suggesting degradation of PE could increase with severity of mutation. As PE has major roles in electron transport chain activity and endoplasmic reticulum homeostasis, the depletion could increase production of reactive oxygen species or unfolded protein response.

Lactic acid and lactone were here seen to increase with *GBA1* variant severity and are related to glucose metabolism and thus could be linked to GCase cleavage. In CSF, lactate is thought to be an indirect measure of brain mitochondria activity and redox balance and therefore a measure of oxidative stress.^20^ Moreover, lactate promotes synthesis of fatty acids^21^ and may be responsible for the observed increase of fatty acyls both by GC-MS and LC-MS. Whether this could represent a marker of increasing disease severity or faster progression warrants investigation in future longitudinal studies.

## Oxidative Stress

Cytochrome P450s are involved in oxidative stress and mitochondrial dysfunction and here the pathway is downregulated in *GBA1*-PD.^22^ Cytochrome P450 enzymes metabolise arachidonic acid to eicosanoids, relating them to dysregulation in the arachidonic acid metabolism pathway.^23^ In earlier work, we have also seen dysregulation of arachidonic acid metabolism pathway between disease and control.^7^ The decrease of an eicosanoid with variant severity may further suggest dysregulation in conversion to arachidonic acid. Additionally, nicotinate and nicotinamide metabolism, which has antioxidant functions was upregulated in iPD. These pathways infer a difference between iPD and *GBA1*-PD in the way the oxidative stress mechanisms occur. There is some evidence *GBA1* variants impair autophagy which in turn reduces oxidative imbalance through sequestered GCase causing an unfolded protein response, which supports this.^24,25^

## Amino Acids and Neurotransmitters

Folate biosynthesis was found to be upregulated in the *GBA1*-PD group. Folate deficiency has previously been found in PD,^26^ with reduced folate metabolism labelled a potential regulator of alpha synuclein toxicity, and indirectly linked with dopaminergic neuronal loss in mouse models.^27,28^ The downregulation in iPD may suggest this mechanism is more pronounced. The GC-MS analysis of serum supports this with decreased dopamine and elevated L-Dopa in iPD, though these metabolites may be directly consequential of administered medication.

Valine, leucine and isoleucine – branch chain amino acids (BCAA) involved in protein biosynthesis – show upregulated synthesis and degradation in iPD., our previous work found the degradation pathway as significant between drug naïve PD and control. In this work, the elevation may impair uptake of aromatic amino acids such as tryptophan due to competition in amino acid transporters.^29^ Reduced BCAAs have been found in plasma of PD patients, as well as decreasing with advancement of PD, suggesting *GBA1*-PD presents as more severe form of PD.^30^

Finally, when comparing drug naïve to medicated participants there are features appearing directly related to administration of l-dopa. The lack of any significant features by t test in sebum could be due to a need for multivariate analysis.

Separation in the scores plot indicates metabolomic changes are present on skin, but perhaps the individual features do not strongly vary alone, and the discrimination is a combination of variance in several features.

For *GBA1*-PD *vs*. iPD, and drug naïve *vs*. medicated PD, GC-MS headspace of sebum shows comparable separation in supervised scores plots compared to GC-MS analysis of serum. This indicates sebum provides an alternative, non-invasive biofluid for classification, though has limitations in detection and identification. For meaningful putative annotations and for pathway analysis, serum and/or LC MS of sebum can provide complementary information.

## Conclusion

We here demonstrate changes in both serum and sebum that differentiate *GBA1*-PD from iPD with high sensitivity and specificity. Analysis of the differences between the classes suggest alterations in lipid regulation, oxidative stress and dopamine pathways that likely reflect the underlying pathogenetic pathways in the PD patients. When features are investigated as a function of severity of *GBA1* variant, several show differential regulation, including sphingolipids, glycerophospholipids, fatty acyls and lactate. Finally, both serum and sebum can differentiate between drug naïve and medicated participants by supervised analysis, with serum revealing significant metabolites including tyrosine and dopamine derivatives. In conclusion, these findings inform on disease mechanisms specific for subtypes of *GBA1*-PD. Future studies should evaluate any association of these markers with disease severity and progression in PD populations, and their potential as markers of disease development within asymptomatic *GBA1* variant carriers.

## Ethics

Participants were recruited via the RAPSODI GD and PD Frontline study, approved by the local Ethics Committees (London – Queen Square REC: 15/LO/1155). All participants signed informed consent upon enrolment.

## Data Statement

Raw data sets generated during the current study are available from MetaboLights Repository https://www.ebi.ac.uk/metabolights/search with the study Identifier MTBLS10743.

## Contributions

AS and PB conceived and supervised the study. CWD extracted and analysed the samples and performed data and pathway analysis. EM performed patient enrolment recruited and collected samples and metadata. CWD led the drafting of the manuscript with contributions from all authors.

## Acknowledgements

This study used instrumentation funded by BBSRC (award BB/L015048/1) and we acknowledge the support of staff in the Mass Spectrometry and Separations Facility of the Faculty of Science and Engineering at the University of Manchester. We are grateful to all the participants who took part in this study as well as PIs and nurses at the recruiting centres. Finally, we thank Waters for their continued support to the Michael Barber Centre for Collaborative Mass Spectrometry.

