## Supplementary Information for "Metabolomic Changes in Idiopathic and *GBA1* Parkinson’s Disease"

SI 1 - Materials and Methods

Sebum was collected from the upper back of participants with medical gauze. Sampled gauze swabs were sealed in plastic bags. Serum samples were obtained from whole blood. All samples were stored at -80 °C until analysis. Samples were blinded and randomised before extraction and analysis.

Serum was thawed on ice. Samples were aliquoted (50 uL) and a portion of each (15 uL) combined in a pooled QC which was subsequently aliquoted after addition of all samples. Ice-cold methanol (200 uL) deproteinized the serum, followed by vortex mixing and centrifugation (15 min, 12000 rpm). Supernatant was transferred to a second tube and dried under vacuum centrifuge (12 hrs, ambient temperature).

LC-MS Analysis

Samples were reconstituted with the addition of 100 uL water, vortex (10 sec), sonicated (10 min), vortex (10 sec) and 80 uL transferred to an LC vial. LC-MS analysis was performed on an Acquity Premier UPLC (Waters) coupled to a ZenoTOF 7600 mass spectrometer (Sciex). An Acquity UPLC BEH C18 Column (1.7 um, 2.1 mm x 100 mm) heated at 50 °C separated the samples. Mobile phase A was water and mobile phase B was methanol both with 0.1% formic acid. An injection volume of 5 uL was used and the flow was at 0.3 mL/min. In both ionisation modes the gradient elution started at 2% B, was held for 1 min then increased to 98% B at 16 min where it was held until 20 mins, the gradient reduced back to 2% B to 22 min to equilibrate the column. Full MS spectra were obtained within the range *m/z* 50-2000, with mass calibration performed every 5 injections by Sciex X500 calibration solution. The source temperature was kept at 400 °C, capillary voltage set to 5500 V in positive ionisation and -4500 V in negative ionisation. In both, IDA mode operated using top 40 with collision energy of 40V (-40V in negative ionisation mode).

GC-MS Serum Analysis

Dried down samples were derivatised by addition of methoxy and trimethylsilyl groups first by addition of O-methoxyamine・HCl in pyridine (50 uL) heating at 65 °C for 40 min followed by 50 uL of MSTFA and heating at 65 °C for 40 min. Samples were centrifuged for 15 min at 12,000 rpm and 100 uL of each transferred to the GC vial.^1^ Derivatised serum samples were analysed on an 7890B GC coupled with a 7200 Mass Spectrometer (both Agilent). Separation was on an VF-5MS (30 m x 120 um x 0.25 um) column with a flow rate of 1 mL/min and a split ratio of 20. The gradient of the oven was 70˚C held for 4 min, then increased at a rate of 14˚C/min until it reached 300˚C where it was held for 4 min giving a total run time of 24.4 min. Full MS Spectra were obtained within the range *m/z* 35-500 and source and transfer temperature held at 230˚C and 280˚C respectively.

For both LC-MS and GC-MS the pooled QC sample was used for analytical reproducibility and filtering of non robust features. QC samples were injected at the beginning of analysis (*n* = 5), after every 5 sample injections and at the end of all samples (*n* = 3).

Headspace GC-MS Sebum Analysis

Sebum was collected from the back of individuals using gauze swabs. Participants were asked to not shower 24 hours prior to sample collection. After collection, samples were stored in 15 mL centrifuge tubes at -80 ˚C until analysis.

The samples were transferred to headspace vials and were analysed on an 7890A GC (Agilent) with a 5975 MSD (Agilent) and equipped with a CTC PAL autosampler. The Vacuum-assisted ITEX technique was used for enrichment, extraction and collection of volatile compounds in the headspace. Samples were incubated at 60˚C for two mins after which the headspace was extracted under vacuum onto the ITEX trap for 15 min with the trap held at at 30 ˚C. The trap contents were desorbed into the GC inlet by heating the trap to 300 ˚C where analytes were re-focussed onto a tenax trap in the inlet at 20 ˚C before increasing the inlet temperatures at a rate of 700 ˚C to 300 ˚C where it was held for 5 min. The GC operated in solvent vent mode. Compounds were separated on an Agilent VF-5MS column (30 m x 250 um x 0.1 um) with a flow of 0.5 mL/min. The oven ramp started at 40 ˚C with a 2.5 min hold before increasing at a rate of 10 ˚C/min until it reached 300 ˚C for a total run time of 28.5 min. The mass selective detector (MSD) operated in scan mode for a mass range of *m/z* 30-500, source temperature was held at 230 ˚C and the quadrupole at 150 ˚C. A five minute solvent delay was used on the MSD. In the absence of a biological QC, an SST of two volatile standards was used (limonene and carvone) and injected throughout.

SI 2 - Data Analysis

LC-MS

The raw LC-MS data files were deconvolved and aligned to the most suitable QC injection in Progenesis QI. Both polarities were subsequently batch corrected using QCs and injection order. Features with > 20% RSD in the QC injections were removed, resulting in 5353 features for positive ionisation and 3575 for negative ionisation. When it was seen the QCs clustered tightly together they were removed from analysis. Putative identifications were assigned using Lipid Maps, Lipid Blast and HMDB at MSI level 2.

GC-MS

The GC-MS data were converted to open source mzXML format^2^, deconvolved using in-house scripts and eRah package in R, then batch corrected using QCs and injection order (for sebum just by injection order).^10^ Identifications at MSI level 2 were assigned by the GOLM database where MatchFactor < 80 were not considered, and in sebum data TMS derivatives were discounted.

For serum the 589 features were refined by removal of those with >20% RSD in pooled QC injections, resulted in 365 features. For all serum data the QC injections clustered tightly in an unsupervised scores plot which was considered satisfactory and thus QC was not analysed further. For sebum the output data matrix had 634 features. Due to absence of pooled QC in sebum, SST were manually overlaid and it was determined there was no significant variation in intensity and retention time. Features with higher average intensity in blank swabs than sample swabs were removed, resulting in 535 features for modelling.

All serum data was log transformed and mean centred scaled, sebum data was additionally normalised to sum to account for different amounts of sebum collected.

Mummichog and Pathway Analysis

LC-MS based metabolomics faces a large bottleneck in identification of features due to richness of data and lack of databases and standards.^5,6^ Mummichog predicts the activity and significance of a metabolite network directly from features, overcoming the need for identifications.^7^ From data obtained of enriched pathways and features contributing to these, we can try and identify the analytes using traditional methods.

For GC-MS, lack of a parent ion means identifications are assigned by matching fragments and fingerprints in the mass spectrum. This, combined with the limits on analytes detected in GC-MS means pathway analysis is instead performed using the putative identifications. Using Metaboanalyst pathway analysis, the identifications and HMDB assignments and Homo sapiens KEGG library we obtained significant pathways. In the case of sebum, as many identifications are alkanes and fatty acids, indicative of lipid breakdown^8^, pathway analysis is not possible. In these cases the identifications of features of interest are described as their class rather than annotation of carbon number, due to lack of information of chain length.

Table S1: Demographics of participants used in the study with significance testing between the groups used in the three models. Age and BMI are notated as mean ± standard deviation, p-values for model 1 and 3 were calculated by student t.test and model 2 by ANOVA. * denotes the parameter was significant between cohorts.

| Model 1 | GBA1-PD (n=25) | iPD (n=25) |  | p-value |
| --- | --- | --- | --- | --- |
| Male:Female | 12:13 | 13:12 |  |  |
| Age | 63.28 ± 5.91 | 62.04 ± 6.31 |  | 0.48 |
| BMI | 25.11± 3.53 | 25.25 ± 3.74 |  | 0.89 |
| Model 2 | Risk (*n=*12) | Mild (*n=*6) | Severe (*n=*7) | *p*-value |
| Male:Female | 7:5 | 3:3 | 2:4 |  |
| Age | 61.25± 5.48 | 66.5±6.35 | 64± 5.66 | 0.20 |
| BMI | 23.01± 2.33 | 23.05 ± 2.32 | 28.63 ± 3.82 | 0.0026* |
| Model 3 | DN (*n=*15) | Med (*n=*15) |  | *p*-value |
| Male:Female | 7:8 | 18:17 |  |  |
| Age | 63.87± 6.12 | 62.14± 6.08 |  | 0.36 |
| BMI | 24.81 ± 3.54 | 25.34± 3.67 |  | 0.64 |

Table S2: Mummichog pathway analysis of samples analysed by LC-MS in positive and negative ionisation comparing the phenotypes GBA and idiopathic PD (idiopathic PD). The features annotated to six metabolic networks displayed higher intensity in the idiopathic PD class and features annotated to three networks displayed higher intensity in the GBA class. Five pathways had multiple features that displayed different regulations, and the sphingolipid metabolism pathway had different features in positive and negative ionisation mode that displayed different regulation.

| Pathway | Regulation |
| --- | --- |
| Terpenoid backbone biosynthesis | Upregulated in iPD |
| Tryptophan metabolism | No regulation |
| Valine, leucine and isoleucine degradation | Upregulated in iPD |
| Valine, leucine and isoleucine biosynthesis | Upregulated in iPD |
| Nicotinate and nicotinamide metabolism | Upregulated in iPD |
| Arachidonic acid metabolism | Upregulated in *GBA1-*PD |
| Fructose and mannose metabolism | No regulation |
| Sphingolipid metabolism | Different regulation between polarities |
| Phenylalanine, tyrosine and tryptophan biosynthesis | No regulation |
| Amino sugar and nucleotide sugar metabolism | Upregulated in iPD |
| Porphyrin metabolism | Upregulated in *GBA1*-PD |
| Phenylalanine metabolism | No regulation |
| Folate biosynthesis | Upregulated in *GBA1*-PD |
| Primary bile acid biosynthesis | No regulation |
| Drug metabolism – cytochrome P450 | Upregulated in iPD |

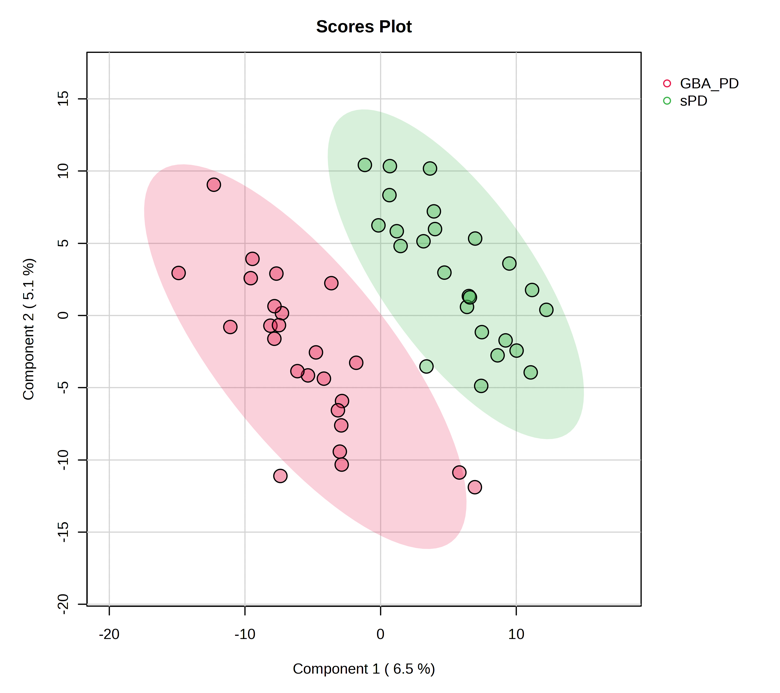

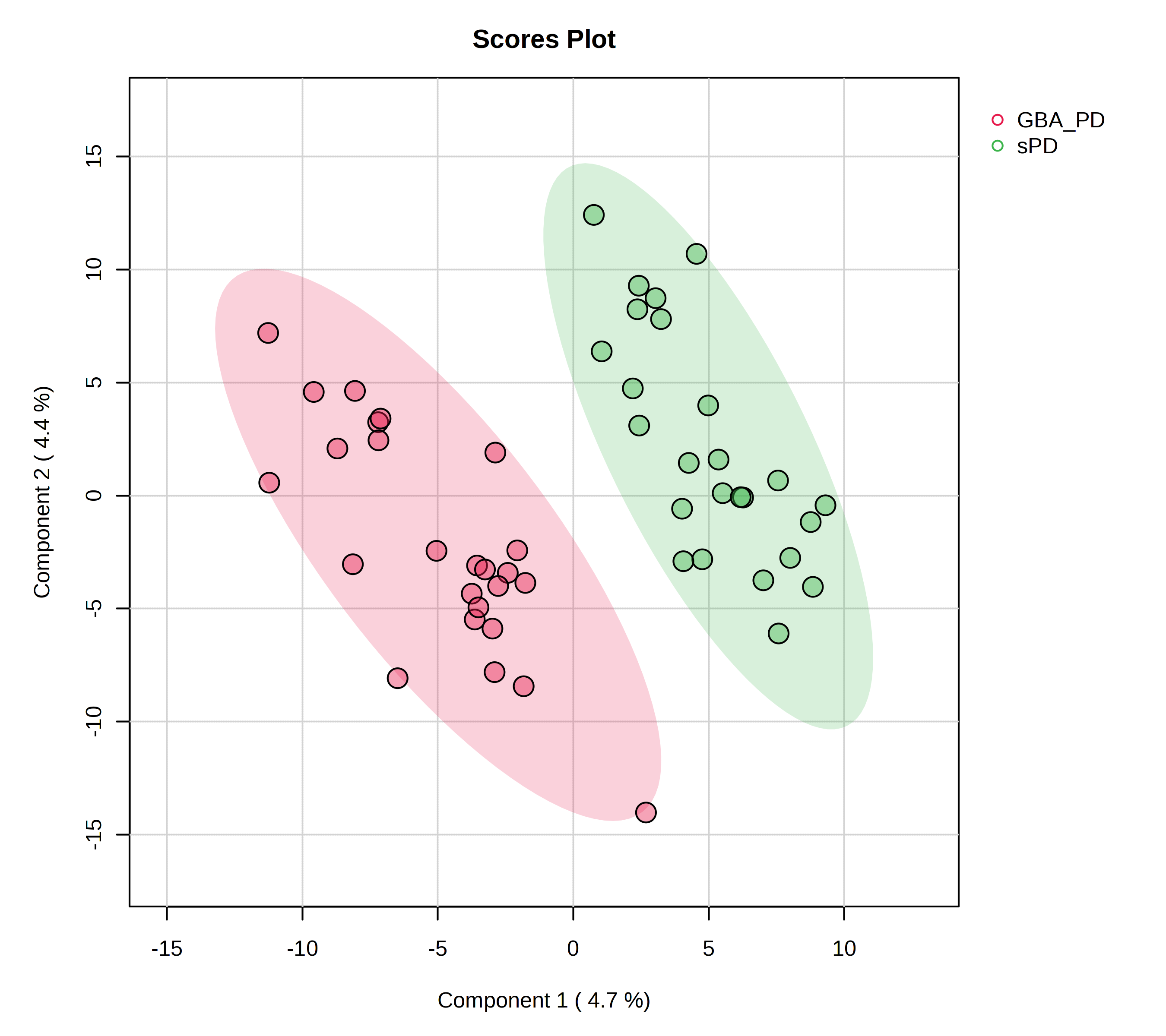

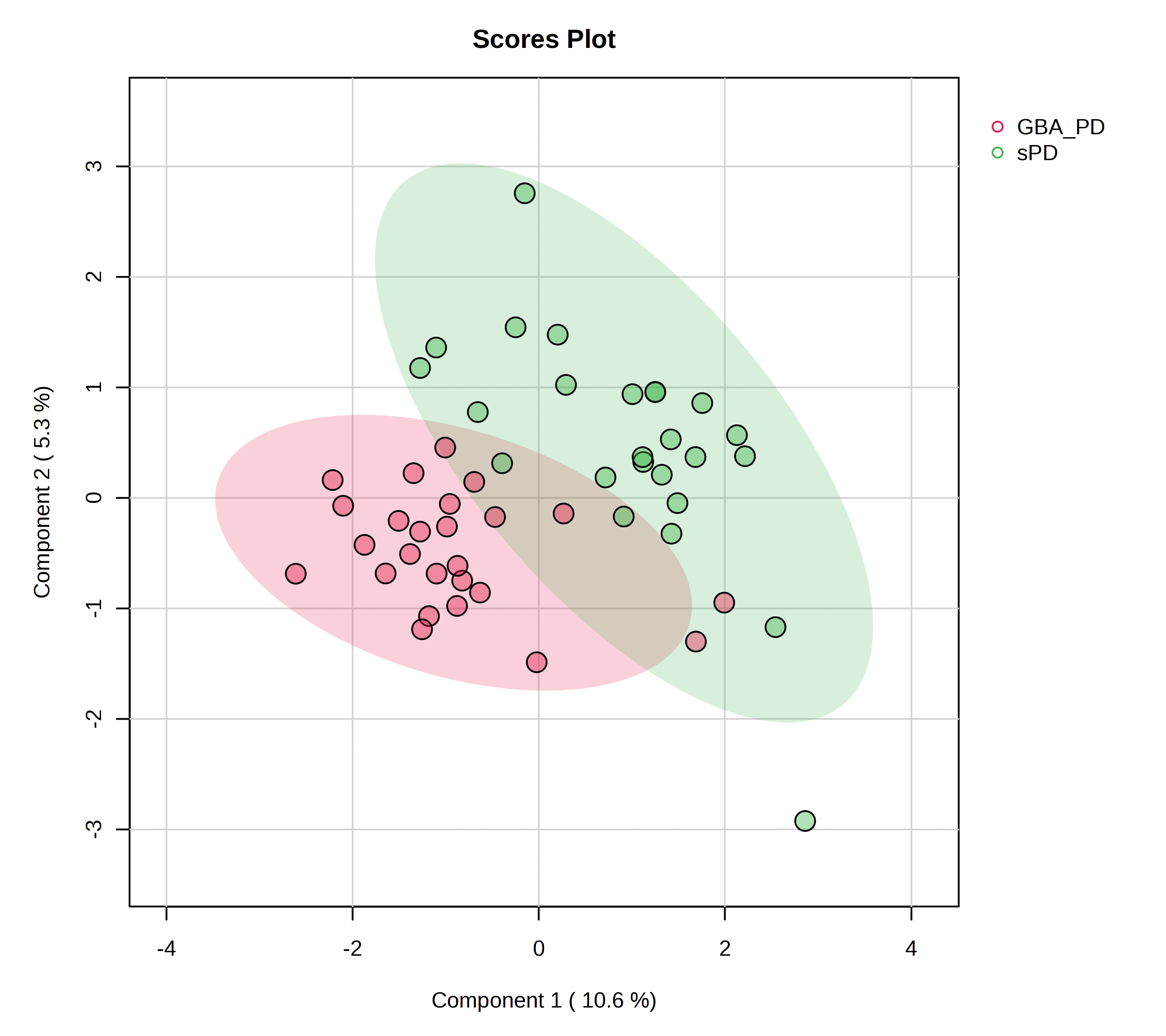

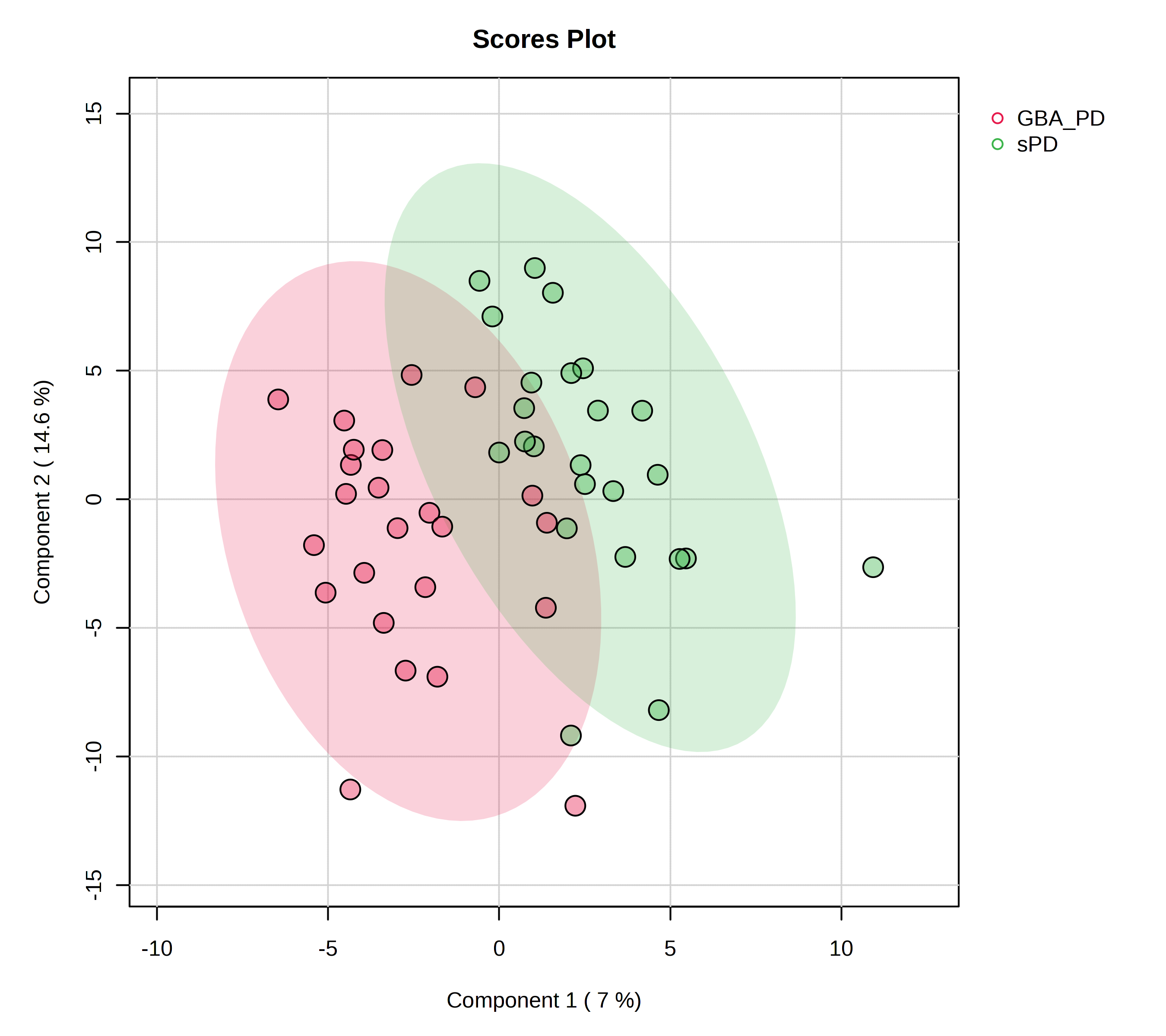

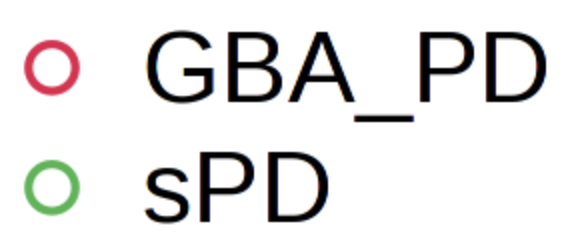

A

B

C

D

Figure S1: PLS-DA supervised scores plots of the GBA1-PD (red) vs. iPD (green) models by all analytical platforms. A) displays the LC-MS in positive ionisation, B) LC-MS in negative ionisation, C) GC-MS of serum and D) Headspace GC-MS of sebum. The LC-MS analyses show complete separation of the sample groups where the GC-MS display clustering of the samples but with some overlap.

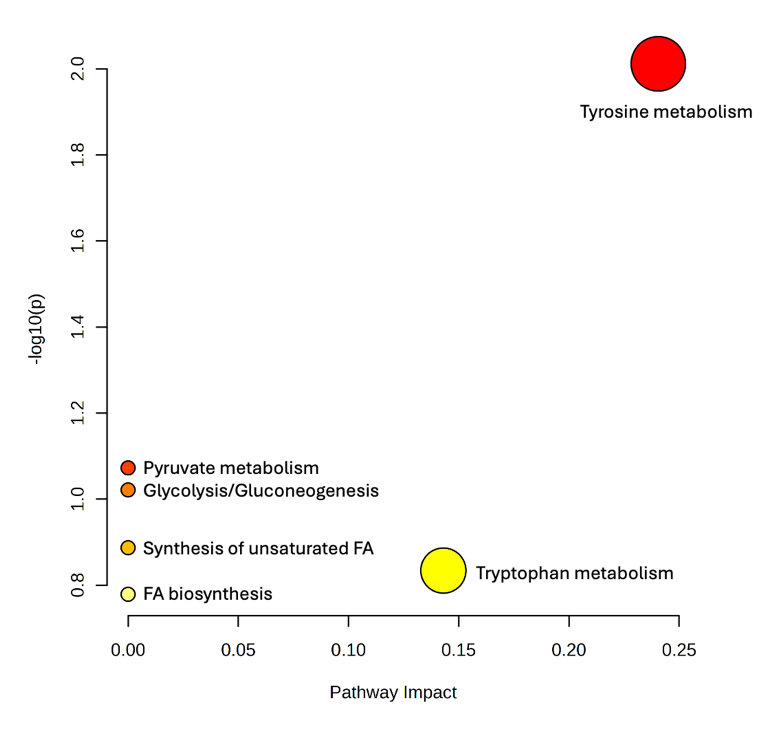

Figure S2: Pathway analysis map of GC-MS serum investigating pathways dysregulation in GBA1-PD vs. iPD. It can be seen that tyrosine metabolism is the only significant pathway with p < 0.05 (0.00973 and -log(p) value 2.012).

Figure S3: PLS-DA supervised scores plots of the Drug naïve (red) vs. medicated (green) models by all analytical platforms. A) displays the LC-MS in positive ionisation, B) LC-MS in negative ionisation, C) GC-MS of serum and D) Headspace GC-MS of sebum. The LC-MS analyses show complete separation of the sample groups where the GC-MS display clustering of the samples but with some overlap.

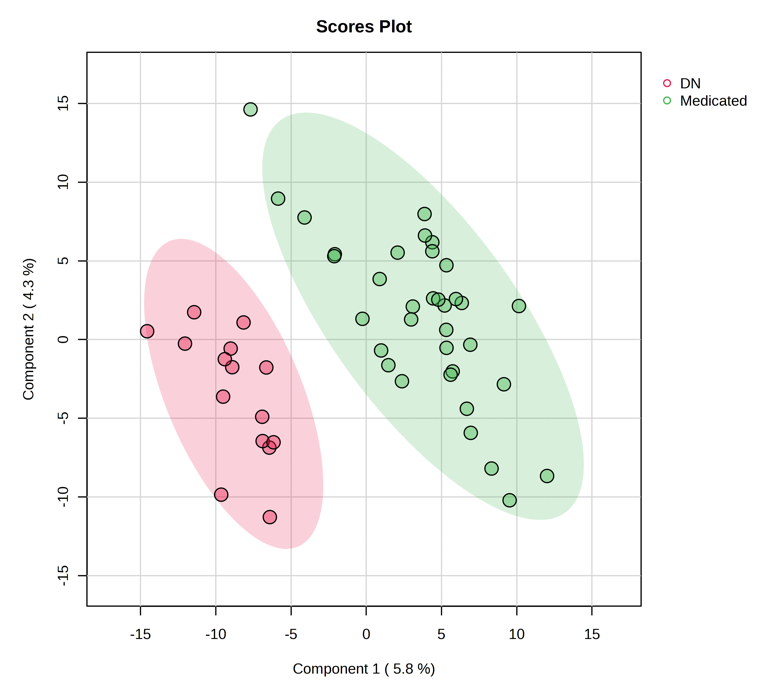

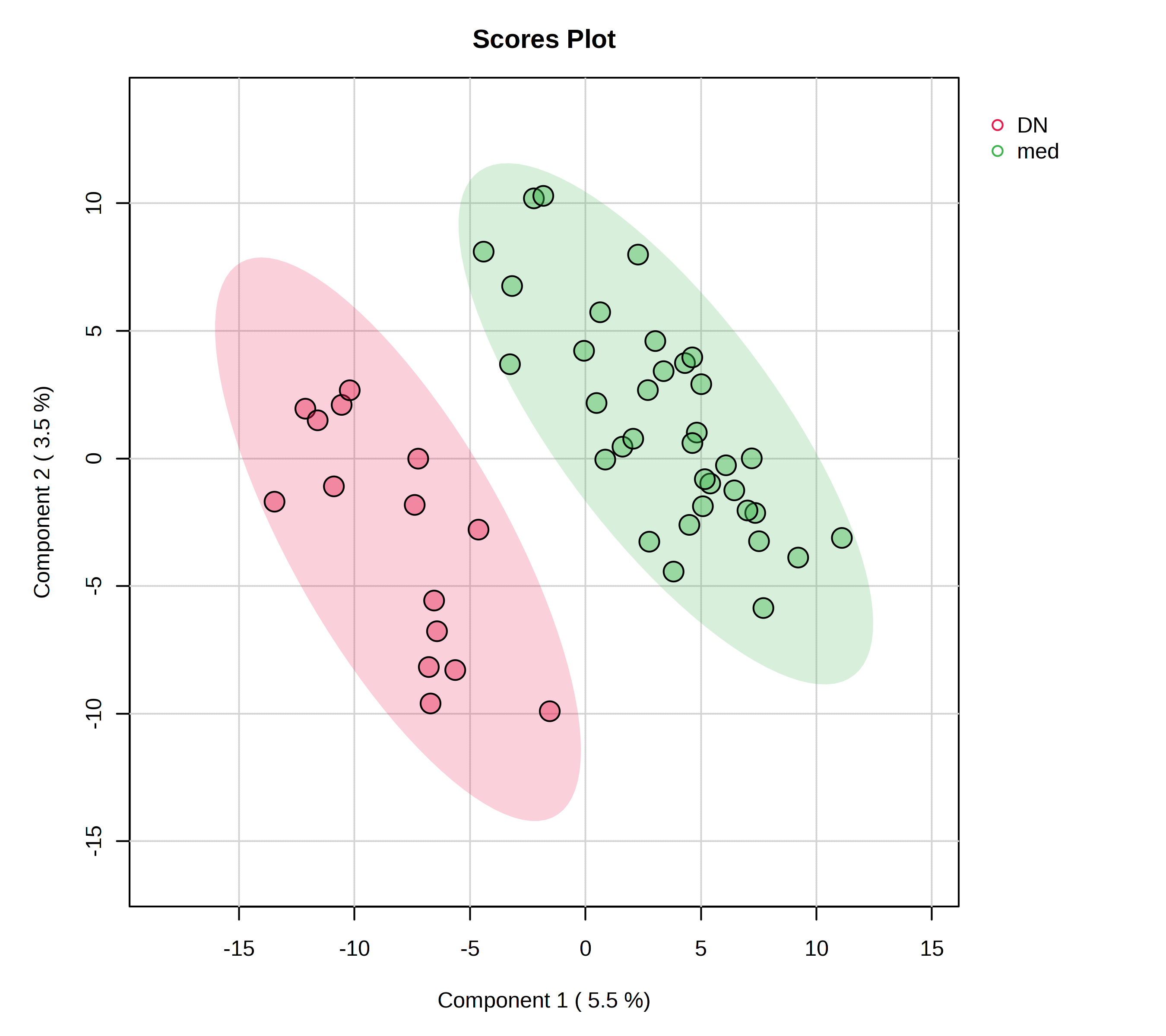

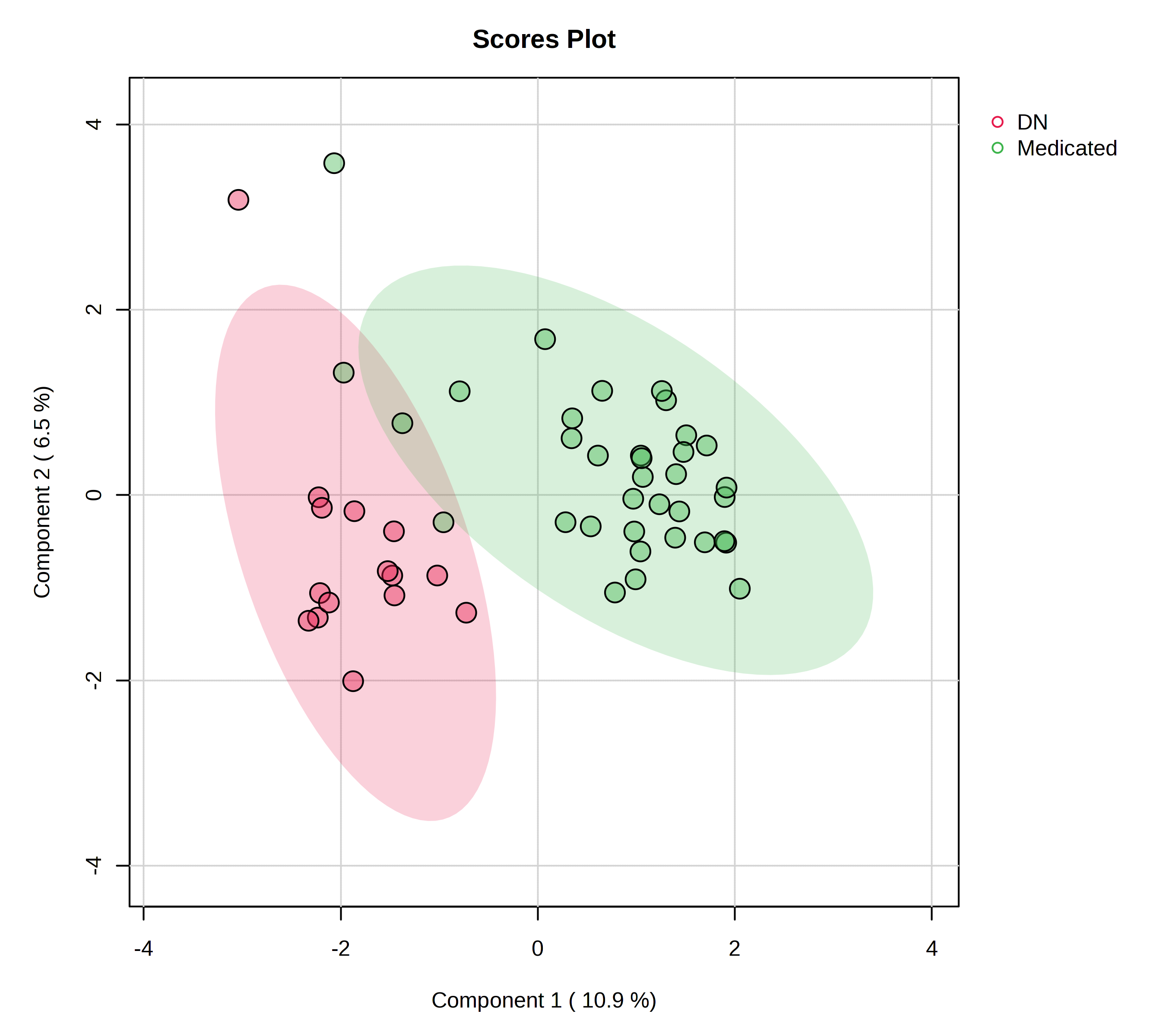

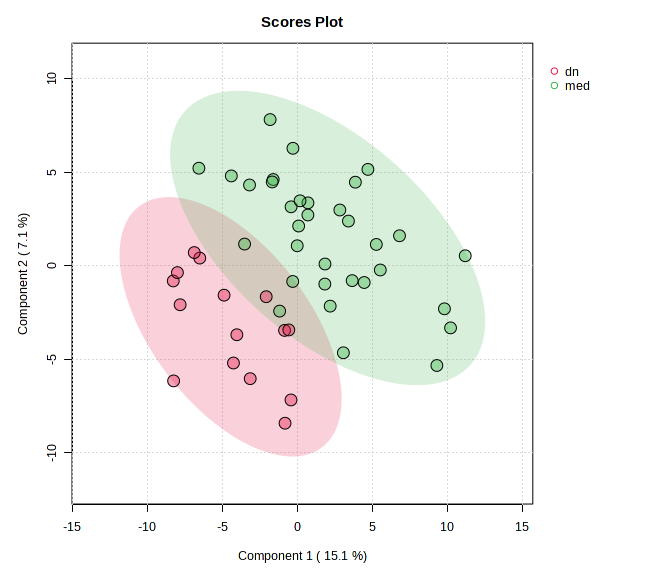

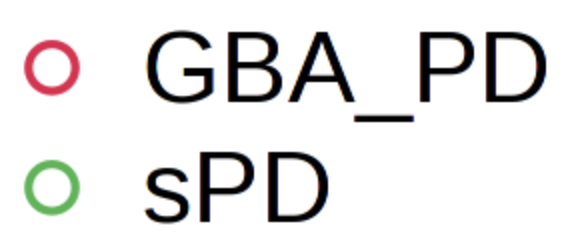

Drug Naive

Medicated

A

B

C

D

Table S3: Medication taken by the five participants of the medicated cohort not currently administered Levodopa.

| Disease Class | Medication |
| --- | --- |
| iPD | Rasagiline |
| iPD | Selegiline |
| *GBA1*-PD | Pramipexole |
| *GBA1*-PD | Trihexyphenidyl, Amantadine, Selegiline, Ropinirole |
| iPD | Previously: Levodopa and Rasagiline |

SI 3 - Pearson’s Correlation

Pearsons correlation was undertaken looking at medication against the feature list for all analysis techniques, a feature was classified as correlated when it had correlation score >0.8 and significance <0.05. By GC-MS and headspace GC-MS, no features were found to show correlation. In LC-MS both ionisation modes found features to be significant, in positive there were eleven features and in negative there were twelve, the features are described below in the table.

Table S4: Features with correlation >0.8 and significance <0.05 using Pearson’s correlation of LC-MS features against medication.

| Feature | Medications | Putative ID | Polarity |
| --- | --- | --- | --- |
| 12.03_331.0253 | Apomorphine Pen, Opicapone, Safinimide | Exogenous | + |
| 9.45_306.1076 | Entacapone | Purine Derivative | + |
| 8.11_302.1423 | Apomorphine Pen, Opicapone, Safinimide |  | + |
| 12.50_528.2795 | Apomorphine Pen, Opicapone | Exogenous | + |
| 7.48_481.1326 | Entacapone |  | + |
| 5.88_134.1093 | Amantadine | Exogenous | + |
| 7.88_481.1326 | Entacapone |  | + |
| 12.92_421.2354 | Apomorphine Pen, Opicapone | Glcerophosphoglycerols | + |
| 11.77_623.3307 | Apomorphine Pen, Opicapone |  | + |
| 12.02_582.2468 | Apomorphine Pen, Opicapone | Bilverdin – Hemoglobin breakdown | + |
| 1.62_216.0087 | L-Dopa |  | + |
| 7.77_223.0068 | Apomorphine Pen, Opicapone | Exogenous | - |
| 7.43_481.1327 | Entacapone |  | - |
| 12.65_511.2531 | Apomorphine Pen, Opicapone | Saccharolipid | - |
| 13.10_493.2431 | Apomorphine Pen, Opicapone |  | - |
| 12.34_509.2381 | Apomorphine Pen, Opicapone |  | - |
| 14.17_333.2061 | Apomorphine Pen, Opicapone | Eicosanoid | - |
| 2.32_340.0345 | L-Dopa | Exogenous | - |
| 17.17_355.2842 | Apomorphine Pen, Opicapone | FA Ester | - |
| 12.46_509.2385 | Apomorphine Pen, Opicapone | Plant metabolite | - |
| 11.03_622.3225 | Amantadine | Bile acid derivative | - |
| 2.32_278.0641 | L-Dopa | Carboxylic acid derivative | - |
| 1.61_232.0282 | L-Dopa | Dopamine Sulphate | - |

Pearsons’ Correlation was further calculated with the medications classed into six groups as follows: Dopamine agonist (ropinirole, pramipexole, rotigotine, apomorphine), COMT inhibitors (entacapone, opicapone), MAO-B inhibitors (rasagiline, selegiline, safinamide), Levodopa, Amantadine and Trihexyphenidyl. In all analysis there were no features correlated to these grouped medications.

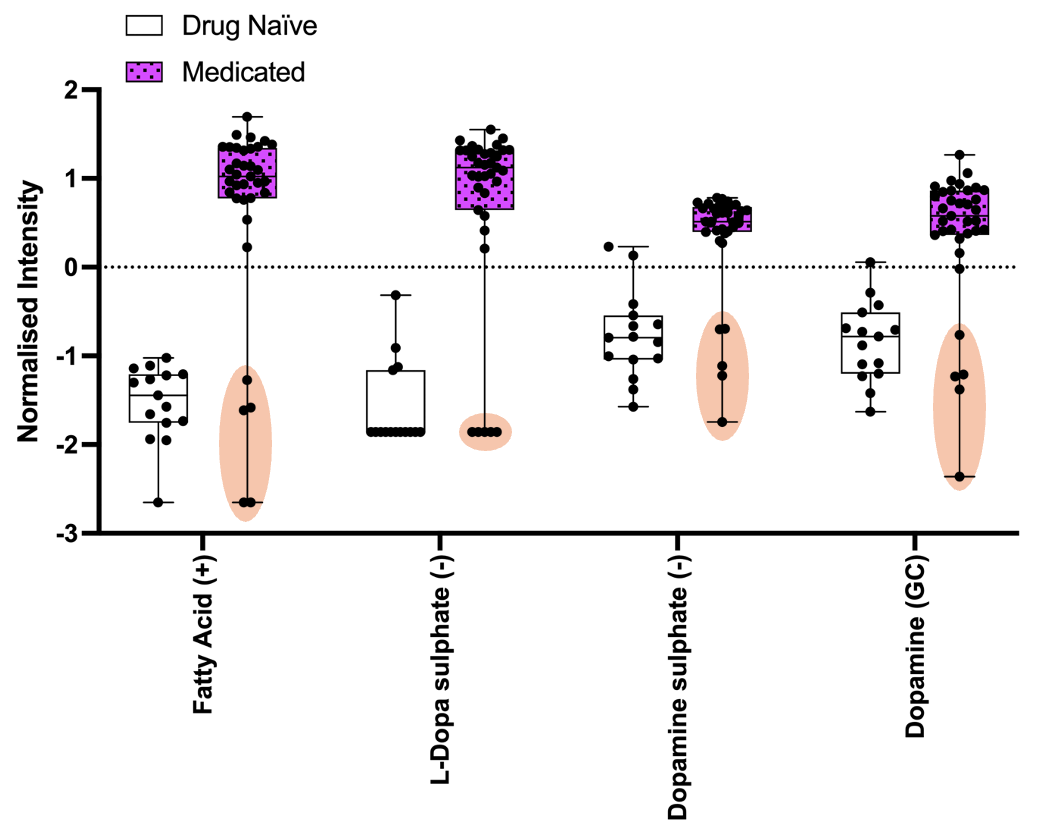

Figure S4: Box and whisker plots for the medicated vs. drug naïve classes displaying five medicated participants as much lower normalised intensity (highlighted in orange ellipses). The five medicated participants with intensity in the region of the drug naïve are the only medicated cohort not receiving l-dopa, and thus it is likely these metabolites are correlated with l-dopa intake. (+) indicates LC-MS positive ionisation (-) is negative ionisation and GC is the GC-MS analysis.
